# Hydrophobic Agent Permeability Assay (HAPA) for Rapid Antibiotic Susceptibility Testing in Gram-Negative Bacteria

**DOI:** 10.1101/2025.05.20.25328050

**Authors:** Yunpeng Guan, Le Zhang, Jian Li, Amy K. Cain, Dan Li, Jiajia Zhou, Lei Jiang, Dayong Jin

**Affiliations:** Institute for Biomedical Materials and Devices (IBMD), University of Technology Sydney, New South Wales, Australia; Infection Program and Department of Microbiology Biomedicine Discovery Institute, Monash University, Victoria, Australia; School of Natural Sciences, Macquarie University, New South Wales, Australia; Zhejiang Provincial Engineering Research Centre for Organelles Diagnostics and Therapy, Eastern Institute of Technology, Ningbo, China

## Abstract

Timely antibiotic susceptibility testing (AST) is essential for effective infection management and resistance control. Standard methods such as broth microdilution require 16-24 hours, delaying appropriate therapy. Existing rapid AST approaches are often limited by high cost, low throughput, or reduced accuracy in heterogeneous bacterial populations. We developed the Hydrophobic Agent Permeability Assay (HAPA), a phenotypic AST method that combines morphology profiling and hydrophobic dye uptake to detect antibiotic-induced membrane changes. HAPA classifies Gram-negative bacterial susceptibility within 4 hours. In susceptible strains, antibiotic-triggered reactive oxygen species (ROS) remodel lipopolysaccharide-mediated membrane permeability, enhancing dye entry. This shift coincides with MIC-level inhibition, enabling accurate breakpoint determination. HAPA demonstrated 97.1% categorical agreement with BMD across 75 clinical isolates, including *E. coli*, *K. pneumoniae*, *A. baumannii*, and *P. aeruginosa*, with error rates below regulatory thresholds. Compatible with automation and point-of-care imaging, HAPA offers a cost-effective, scalable solution for rapid diagnostics and improved antimicrobial stewardship.

## Introduction

Antimicrobial resistance (AMR) is a leading global health challenge, with an estimated 4.7 million deaths associated with bacterial AMR in 2021, disproportionately affecting low- and middle-income countries.^1^ Multidrug-resistant Gram-negative bacteria (MDR-GNB) are particularly concerning due to their increasing resistance to last-resort agents such as carbapenems, polymyxins, and novel β-lactam/β-lactamase inhibitor combinations.^2^ The World Health Organization (WHO) has classified several MDR-GNB under its “Priority Pathogens List”, identifying “ESAKPE” pathogens *Escherichia coli*, *Klebsiella pneumoniae*, *Acinetobacter baumannii*, and *Pseudomonas aeruginosa*, as critical priority pathogens.^3^

While global efforts to combat AMR have focused on the development of new antimicrobials and non-traditional therapies, diagnostic innovation particularly in antibiotic susceptibility testing (AST) has emerged as an equally critical priority.^4^ Timely and accurate AST is essential for enabling appropriate therapy, minimizing empirical broad-spectrum antibiotic use, and reducing AMR selection pressure.^5,6^ However, conventional AST methods such as broth microdilution (BMD), disk diffusion, and agar dilution typically require 16-24 hours to deliver results, delaying effective treatment and contributing to poorer clinical outcomes, particularly in time-sensitive conditions such as sepsis, pneumonia, and bloodstream infections.^7^ In the Intensive Care Unit (ICU), each hour of delayed effective therapy can increase mortality risk by up to 7.6%.^7^

In response, several rapid AST technologies have been developed, including genotypic methods such as polymerase chain reaction (PCR) and whole-genome sequencing (WGS), and phenotypic approaches based on microfluidics, dynamic imaging, or optical readouts.^8^ While genotypic methods offer speed and specificity, they are limited by their inability to detect phenotypic resistance mediated by unknown or complex mechanisms.^9^ In contrast, phenotypic rapid AST methods provide a direct measure of bacterial response to antibiotics but often focus on classifying susceptibility categories, such as “susceptible” or “resistant”, rather than determining MIC breakpoints,^4^ which are the cornerstone of dose selection, pharmacodynamic modelling, and treatment adjustment.^10^ Moreover, many phenotypic platforms rely on expensive instrumentation, limited throughput, or complex microfabrication, restricting their clinical scalability.^4^

To address these limitations, we developed an image-based phenotypic platform that rapidly estimates MIC values by detecting early and antibiotic-induced changes in membrane permeability and morphology. Readouts are both biologically relevant and technically scalable. In this study, we describe the Hydrophobic Agent Permeability Assay (HAPA), which leverages antibiotic-induced oxidative stress and outer membrane remodelling in Gram-negative bacteria to detect susceptibility signatures within 4 hours. By integrating fluorescence imaging with deep learning-based interpretation, HAPA offers a standardized, rapid, and interpretable approach to phenotypic MIC determination, positioning it as a highly promising tool for clinical implementation in time-critical antimicrobial decision-making.

## Materials and Methods

### Bacterial strains and antibiotics

Type strains of *E. coli* MG1655, *P. aeruginosa* ATCC 47085, *K. pneumoniae* 52145, *A. baumannii* ATCC 19606, and *Staphylococcus aureus* ATCC 29213 were used for mechanistic studies. *E. coli* ATCC 25922, *P. aeruginosa* ATCC 27853, *K. pneumoniae* 52145, and *A. baumannii* ATCC 19606 were used for the comparison of antibiotic susceptibility testing. Clinical isolates of *E. coli* (n=13), *K. pneumoniae* (n=15), *A. baumannii* (n=17), and *P. aeruginosa* (n=30) were collected from various diagnostic microbiology laboratories for the purposes of diagnostic testing and database curation^11^. Bacterial cultures were grown overnight in Luria-Bertani Broth (LB) at 37°C with shaking at 250 rpm prior to antibiotic exposure. Stock solutions of ampicillin, cefepime, cefotaxime, meropenem, gentamicin, and ciprofloxacin were prepared in sterile Milli-Q water and filtered through 0.22 µm syringe filters prior to use. Chloramphenicol stock (50 mg/mL) was prepared in 95% ethanol. All antibiotic stocks were aliquoted and stored at −20°C for up to 3 months, or at 4°C for short-term use (≤1 week), and protected from light.

### Antibiotic susceptibility testing and MIC determination

Minimum inhibitory concentrations (MICs) were determined using standard BMD method according to CLSI guidelines (2023 edition).^12^ Approx. 10^4^ cells from an overnight culture were inoculated into each well in a 96 wells plate, and each column was filled with 100 µL fresh Mueller-Hinton Broth (MHB) supplemented with increasing amounts of antibiotics (two-fold serial dilutions). Cells were then incubated in a plate reader at 37°C with shaking at 250 rpm for 18-24 hours. The value of OD_600_ was measured programmatically each hour. MICs were defined as the lowest concentration inhibiting visible growth.

### Nile Red staining and LPS perturbation

Nile Red (Sigma-Aldrich) was used as a hydrophobic membrane-permeable agent.^13^ A 200× stock solution (500 µg/mL) was freshly prepared in DMSO and stored protected from light. For Nile Red staining, unless otherwise specified, bacterial strains were cultured overnight in 5 mL of LB broth at 37°C with shaking at 250 rpm. Cultures were then diluted 1:100 into fresh 5 mL LB and grown under the same conditions until reaching mid-log phase (OD_600_ at 0.4). For antibiotic treatment, 100 µL of this culture was transferred into 10 mL LB medium supplemented with increasing amounts of antibiotics (two-fold serial dilutions). After 4 hours of incubation at 37°C with shaking at 250 rpm, cultures were centrifuged at 4,000 g for 10 minutes. Pellets were washed twice with PBS and resuspended in PBS containing 2.5 µg/mL Nile Red. The suspension was incubated at room temperature in the dark for 30 minutes without shaking.

LPS disruption was induced using either polymyxin B (1-10 µg/mL) or EDTA (1-5 mM). For positive control, *S. aureus* ATCC 29213 was included as a Gram-positive comparator.

For microscopy, agarose pads were prepared by pipetting 30 µL of freshly prepared 1.5% (w/v) agarose in PBS onto a microscope slide, followed by immediate placement of a coverslip.^14^ After solidification (≥15 minutes), the coverslip was gently removed. Then, 2 µL of the stained cell suspension was placed on the agarose pad and covered with a new coverslip for imaging.

### Microscopy and image acquisition

Bright-field and fluorescence images were acquired using a spinning disk confocal microscope (Evident, Australia) equipped with a 100× oil immersion objective (NA 1.45) and a scientific sCMOS camera (Prime 95B, TVS). Nile Red fluorescence was excited using a 561 nm laser, and emission was collected through a 617/73 nm bandpass filter. To ensure reproducibility, laser power was maintained at 40% across all imaging sessions. For each field of view, a z-stack of 15 optical sections was acquired at 0.1 µm intervals with 100 ms exposure per frame. ImageJ was used for preliminary visualization, and raw TIFF files were retained for downstream quantitative analysis.

### Plasmid transformation

Chemically competent *E. coli* MG1655 wild-type cells were prepared using a standard calcium chloride method.^15^ For each transformation, 50 ng of purified plasmid DNA (Table 1) was added to 100 µL of competent cells and incubated on ice for 30 minutes, followed by heat shock at 42 °C for 90 seconds. Transformed cells were recovered in 500 µL of LB medium at 37 °C with shaking for 1 hour. Cells were then plated onto LB agar plates containing the appropriate antibiotic for plasmid selection: 50 µg/mL ampicillin and 50 µg/mL kanamycin for WT-pKD4. Plates were incubated overnight at 37 °C before downstream experiments.

**Table 1.** Plasmid used in this study.

| Plasmid | Features | Source |
| --- | --- | --- |
| pKD4 | Non-replicating plasmid used for gene deletion via $\lambda$ Red recombination; contains a kanamycin resistance cassette flanked by FRT sites and an ampicillin resistance marker for plasmid selection. | Addgene |

### Measurement of ROS levels

Intracellular ROS levels were measured using the Cellular ROS Assay Kit (Abcam, ab113851) following the manufacturer’s protocol. Overnight bacterial cultures were grown in LB medium, then treated with antibiotics (1× MIC) at 37°C for 2 or 4 hours with shaking at 250 rpm. After treatment, cells were washed twice with 1× assay buffer to remove residual antibiotics and debris and subsequently resuspended in fresh 1× buffer. Cells were then incubated with the fluorescent ROS probe DCFDA (2′,7′–dichlorofluorescin diacetate; final concentration: 10 µM) for 30 minutes at 37°C in the dark to prevent probe degradation. After incubation, 500 µL of the stained cell suspension was immediately analysed using a CytoFLEX LX flow cytometer (Beckman Coulter). Fluorescence was detected using the FITC channel (excitation: 488 nm; emission: 525 nm). Data were analysed using FlowJo software (Version X, Tree Star Inc.). Unstained controls and untreated antibiotic-negative controls were included to confirm probe specificity and establish baseline fluorescence.

### Hydrophobic Agent Permeability Assay (HAPA)

HAPA was designed as a rapid, single-cell-level phenotypic assay to estimate antibiotic susceptibility by detecting membrane permeability and morphological changes. The procedure consists of three main steps. First, bacterial cultures were treated with a two-fold serial dilution series of antibiotics for 4 hours at 37°C with shaking. The concentration range for each antibiotic spanned below and above the corresponding CLSI MIC breakpoints to capture the full phenotypic response profile. Second, cells were stained with 2.5 µg/mL Nile Red for 30 minutes at room temperature in the dark. After staining, samples were immediately imaged using a high-resolution fluorescence microscope. Third, fluorescence and morphological features were analysed at the single-cell level. Phenotypic classification based on predefined fluorescence and morphological response patterns at the single-cell level. Susceptibility is determined by comparing observed phenotypes to MIC breakpoint-associated profiles previously benchmarked against CLSI standards.

### Deep-learning model for automated classification

Brightfield images were first pre-processed by applying contrast stretching and Gaussian filtering to enhance bacterial boundary visibility and reduce noise. The pre-processed images were then used to train a U-Net convolutional neural network for bacterial segmentation, supervised using manually annotated cell masks. The segmentation loss function was defined as a weighted combination of Dice Loss, Intersection over Union (IoU) Loss, and Binary Cross-Entropy (BCE) Loss to balance pixel-wise accuracy and overlap-based segmentation quality. The U-Net model was trained using the Adam optimizer (learning rate 0.00001) for 200 epochs. Segmentation performance was evaluated by Accuracy, Mean Overlap Coefficient (MOC), Recall, and Precision, all exceeding 90% on the validation set. Segmentation masks were further manually inspected and corrected to ensure accurate delineation of bacterial regions. These masks were then aligned with corresponding fluorescence images to extract fluorescence features restricted to bacterial areas. For each masked fluorescence image, multiple features were calculated, including the fluorescence-positive pixel ratio, mean fluorescence intensity, sum of fluorescence intensity, maximum fluorescence intensity, bright area proportion, number of bright spots, mean area of bright spots, and standard deviation of fluorescence intensity. The extracted fluorescence features, together with the masked fluorescence image patches, were used as inputs to a second U-Net model designed for antibiotic susceptibility prediction. During classification, the fluorescence image was processed through the U-Net encoder, after which the encoded feature map was flattened and concatenated with the extracted feature vector. This combined vector was passed through fully connected layers and the decoder to predict susceptibility status. The classification model was optimized using the BCE loss function. Classification performance was assessed by Accuracy, Precision, Recall, F1 score, and the area under the receiver operating characteristic curve (AUROC). All models were implemented in PyTorch. A batch size of 10 was used for training, with input images resized to 256×256 pixels. Training and evaluation were performed on a workstation equipped with an NVIDIA RTX 4090 GPU.

### Clinical validation and performance metrics

The diagnostic accuracy of HAPA and AI-assisted readout was evaluated by comparing with BMD-based classification. Category agreement, minor errors (mE), major errors (ME), and very major errors (VME) were calculated in accordance with FDA guidance for AST system evaluation. A categorical agreement ≥90%, VME ≤3%, and ME ≤3% were considered acceptable.^16^

### Receiver Operating Characteristic (ROC) analysis

To evaluate the performance of the HAPA assay in predicting antibiotic susceptibility, ROC curve analysis was conducted using the CLSI-defined BMD results as the reference standard. For each isolate-antibiotic pair, the fluorescence-positive cell ratio was calculated as the proportion of Nile Red-stained cells among morphology-filtered cells in a single imaging field. Isolates were classified as susceptible or resistant according to CLSI breakpoints; intermediate isolates were excluded from ROC analysis. Fluorescence-positive ratios were plotted against binary susceptibility outcomes (susceptible = 1, resistant = 0) to generate ROC curves. The area under the curve (AUC) was calculated to quantify the discriminatory power of the assay. AUC values approaching 1.0 indicate strong agreement between the HAPA assay and the gold-standard classification. Statistical significance was assessed using the DeLong test. ROC analyses were performed using GraphPad Prism version 9.0.0 (GraphPad Software, USA).

### Statistical analysis

Statistical analysis was performed using GraphPad Prism v.9.0.0. All data are presented as individual values and mean or mean ± SEM. One-way ANOVA and a two-tailed unpaired Student’s *t*-test using a 95% confidence interval were used to evaluate the difference between groups. A probability value of *P* < 0.05 was considered significant. Statistical significance is indicated in each figure.

## Results

Lipopolysaccharide (LPS), a crucial constituent of Gram-negative outer membranes, restricts the entry of hydrophobic agents due to its tightly packed lipid A domain and dense polysaccharide chains (Fig. 1A).^17^ Nile Red, a hydrophobic agent with environment-sensitive fluorescence,^13^ failed to stain live *E. coli* and *P. aeruginosa*, yet labelled fixed cells (Fig. 1B), indicating exclusion by intact LPS. In contrast, an LPS-deficient *A. baumannii* mutant (*lpxA* frameshift) exhibited markedly enhanced Nile Red staining (Fig. 1C). Similarly, polymyxin B (PMB) or ethylenediaminetetraacetic acid (EDTA) treatment, known to disrupt LPS integrity, significantly increased Nile Red uptake across all tested Gram-negative species in a dose-dependent manner (Fig. 1D-G; Figure S1A and B). Notably, the hydrophilic agent crystal violet displayed uniform staining regardless of LPS presence or concentration (Fig. 1H and I). As a control, Gram-positive *S. aureus* was readily stained by Nile Red (Fig. 1J). These results confirm LPS as a critical barrier against hydrophobic agents, while hydrophilic agents permeate freely (Fig. 1A).

**Figure 1.**
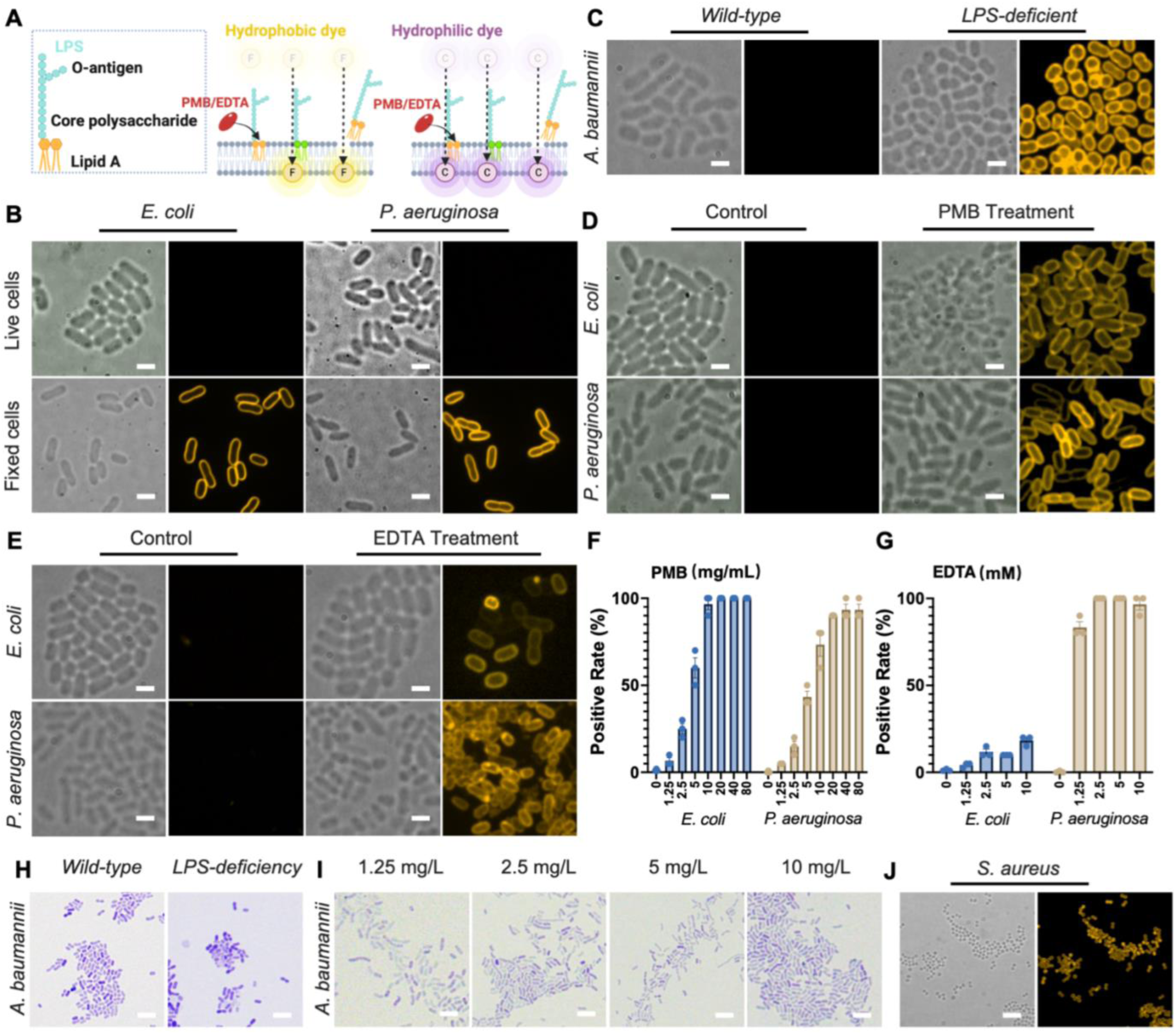
LPS restricts the uptake of hydrophobic agents in Gram-negative bacteria. **(A)** Schematic illustration of the Gram-negative outer membrane highlighting the LPS structure as a selective barrier composed of hydrophilic polysaccharide chains and a lipid A domain, which impedes the entry of hydrophobic agents such as Nile Red **(B)** Nile Red staining of live versus fixed *E. coli* MG1655 and *P. aeruginosa* ATCC 47085 cells. **(C)** Representative fluorescence images showing enhanced Nile Red staining in an *A. baumannii* 19606 *lpxA* mutant lacking LPS, compared to wild-type cells. **(D-G)** Nile Red staining intensity in *E. coli* MG1655 and *P. aeruginosa* ATCC 47085 following treatment with PMB (80 mg/mL) or EDTA (10mM). The positive rate was defined as the proportion of Nile Red stained cells relative to the total number of cells within a single image frame. **(H-I)** Crystal violet staining of wild-type and LPS-deficient *A. baumannii* 19606 strains showed no difference in dye uptake, regardless of LPS presence or treatment with increased concentration of crystal violet. **(J)** *S. aureus* ATCC 29213, a Gram-positive species lacking LPS, was readily stained by Nile Red. Quantification of fluorescence intensity is shown as mean ± SEM from three independent experiments. Scale bars = 1 µm (C-E), 5 µm (H-J).

In addition to Nile Red, we also evaluated other fluorescent agents with varying hydrophobicity, including Nile Blue A, DIOC2(3) (DiD), FM4-64, and BODIPY, to further assess their outer membrane permeability profiles (Fig. S2A). Among the tested agents, Nile Red, Nile Blue A, and BODIPY failed to stain live wild-type Gram-negative bacteria but showed restored uptake in LPS-deficient *A. baumannii*, consistent with selective exclusion by an intact outer membrane. Notably, while Nile Blue A and BODIPY produced weaker fluorescence and higher background signal, Nile Red demonstrated a superior signal-to-noise ratio, making it more suitable for cell imaging of membrane permeability. FM4-64 readily stained both wild-type and LPS-deficient strains, while DiD showed minimal uptake under all conditions. These comparisons led to the use of Nile Red in this study as the representative hydrophobic agent for assessing outer membrane permeability.

To investigate whether antibiotics that do not directly target LPS also modulate outer membrane permeability, we treated *E. coli* and *P. aeruginosa* with cefepime (β-lactam),^18^ ciprofloxacin (fluoroquinolone),^19^ and gentamycin (aminoglycoside)^20^ for 2 and 4 hours. Despite their distinct primary targets, cell wall, DNA replication, and protein synthesis, respectively, all three antibiotics significantly enhanced Nile Red staining in both species after 4 hours of treatment (Fig. 2A). This observation suggests a common downstream effect on membrane properties.

**Figure 2.**
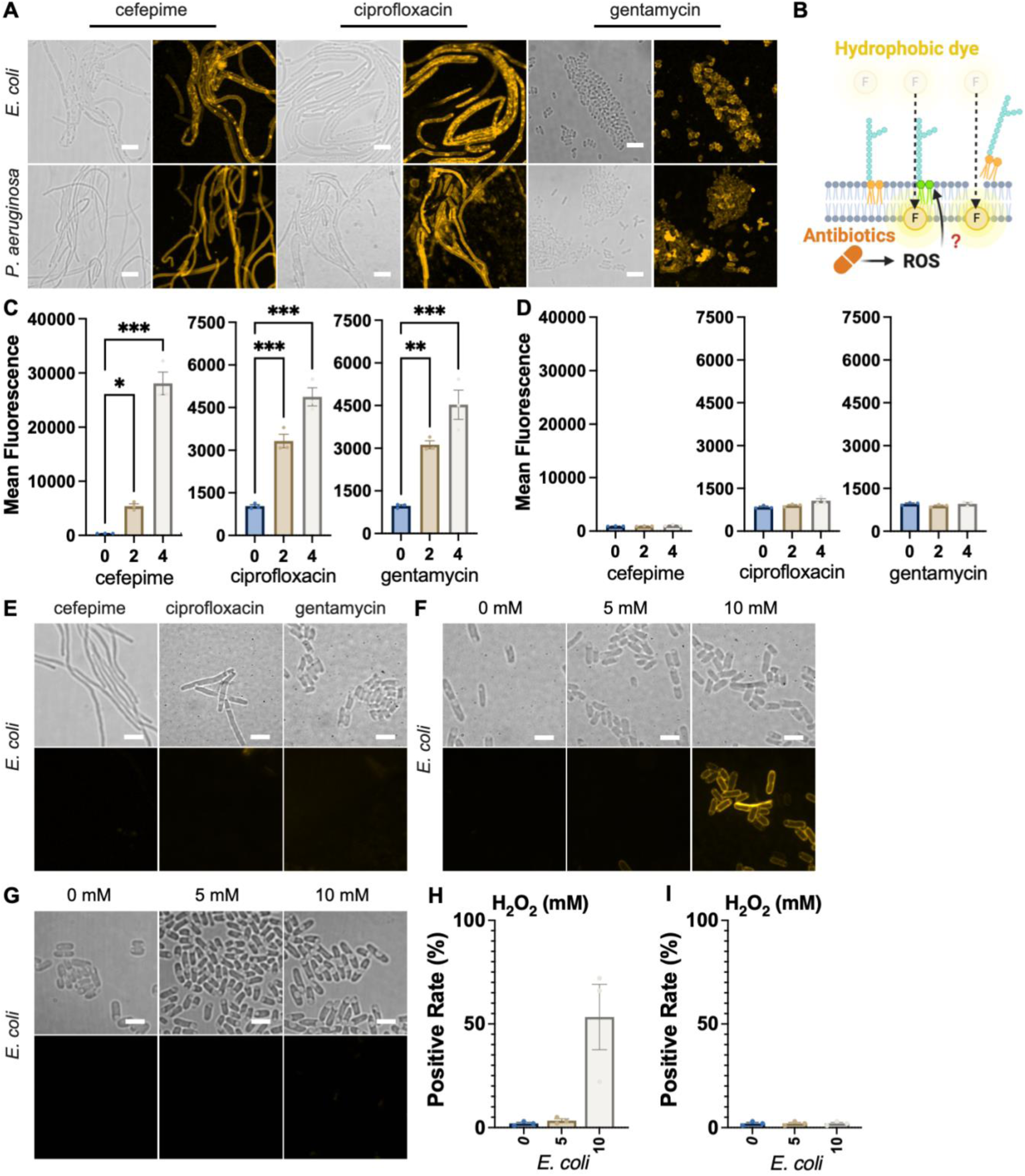
Antibiotic-induced oxidative stress enhances membrane permeability. **(A)** Representative fluorescence and bright-field images of *E. coli* MG1655 and *P. aeruginosa* ATCC 47085 treated with cefepime (0.03 µg/mL for *E. coli* and 1.25 µg/mL for *P. aeruginosa*), ciprofloxacin (0.0125 µg/mL for *E. coli* and 0.5 µg/mL for *P. aeruginosa*), or gentamycin (1 µg/mL for *E. coli* and *P. aeruginosa*) for 4 hours. **(B)** Proposed model: bactericidal antibiotics trigger intracellular ROS production, potentially leading to oxidative modification of lipid A within LPS, increasing its hydrophobicity and allowing enhanced penetration of hydrophobic agents such as Nile Red. **(C)** Intracellular ROS levels measured after antibiotic treatment in *E. coli* MG1655 treated with antibiotics over 0, 2, and 4 hours. **(D)** Co-treatment with GSH (50 mM) abolished antibiotic-induced ROS production in *E. coli* MG1655. **(E)** Representative fluorescence and bright-field images of *E. coli* MG1655 co-treated with antibiotic (cefepime at 0.03 µg/mL, ciprofloxacin at 0.0125 µg/mL, or gentamycin at 1 µg/mL) and GSH (50 mM) for 4 hours. **(F)** Exogenous ROS application via H₂O₂ treatment (0, 5, 10 mM, 1 hour) induced dose-dependent increases in Nile Red fluorescence in *E. coli* MG1655. **(G)** PI staining following H₂O₂ exposure (0, 5, 10 mM, 1 hour) did not show increased signal in *E. coli* MG1655. **(H and I)** Percentage of Nile Red-positive and PI-positive cells in *E. coli* MG1655 after H₂O₂ treatment. The positive rate was defined as the proportion of Nile Red- or PI-stained cells relative to the total number of cells within a single image frame. Quantification of fluorescence intensity is shown as mean ± SEM from three independent experiments. Statistical significance determined by one-way ANOVA; *, *P* < 0.05; **, *P* < 0.01; ***, *P* < 0.001. Scale bars = 5 µm (A), 2 µm (E-G).

A growing body of evidence indicates that diverse bactericidal antibiotics stimulate intracellular reactive oxygen species (ROS) production.^21,22,23^ In eukaryotic cells, ROS induce lipid peroxidation, increasing membrane permeability without necessarily causing structural rupture.^24^ We hypothesised that oxidative stress may alter the outer membrane, potentially via lipid oxidation, thereby increasing hydrophobic agents’ accessibility (Fig. 2B). Consistent with this, we observed a time-dependent increase in intracellular ROS following antibiotic treatment in *E. coli* and *P. aeruginosa* (Fig. 2C; Fig. S3A). Co-treatment with the ROS scavenger glutathione (GSH) suppressed both ROS accumulation (Fig. 2D; Fig. S3B) and the enhanced Nile Red staining (Fig. 2E; Fig. S3C), indicating that ROS generation mediates the increased membrane permeability. To further support the ROS-dependence of this effect, we examined chloramphenicol, a bacteriostatic antibiotic known to inhibit protein synthesis without inducing oxidative stress.^22^ Unlike bactericidal agents, chloramphenicol failed to induce ROS accumulation or enhance Nile Red staining in either *E. coli* or *P. aeruginosa* (Fig. S4A and B), further confirming the role of ROS in modulating outer membrane permeability.

To assess whether ROS is sufficient to alter outer membrane permeability, we exposed *E. coli* and *P. aeruginosa* to increasing concentrations of hydrogen peroxide (H₂O₂). Nile Red fluorescence intensity increased in a concentration-dependent manner (Fig. 2F and H; Fig. S3D and F), supporting a direct role for ROS in modulating membrane accessibility. However, Propidium Iodide (PI) staining, which indicates gross membrane disruption, remained unchanged under the same conditions (Fig. 2G and I; Fig. S3E and G). To further confirm that increased Nile Red uptake does not reflect compromised membrane integrity, we compared PI and Nile Red staining in LPS-deficient *A. baumannii*. While Nile Red readily stained this mutant, PI signal remained absent (Fig. S5A and B), indicating that outer membrane permeability was enhanced without loss of membrane barrier function. Collectively, these findings support a mechanism in which antibiotic-induced oxidative stress alters outer membrane permeability through physicochemical modifications of surface lipid components.

To determine whether this ROS-dependent permeability response is absent in resistant strains, we engineered *E. coli* MG1655 to carry plasmids conferring resistance to either ampicillin or kanamycin. Upon exposure to the corresponding antibiotics for 4 hours, in contrast to their drug-susceptible counterparts (Fig. S6A), these resistant derivatives did not exhibit increased Nile Red staining (Fig. S6B). Furthermore, ROS levels measured after antibiotic treatment for 0, 2, and 4 hours remained unchanged in resistant strains (Fig. S6C and D), indicating a lack of oxidative stress induction. These findings demonstrate that antibiotic-induced ROS production and the associated permeability enhancement occur selectively in susceptible bacteria and are not elicited in resistant strains under identical treatment conditions.

Having established that fluorescent agent uptake enhancement and ROS generation are hallmarks of antibiotic susceptibility, we next investigated whether this phenotypic response could be used to approximate MIC thresholds across a range of antibiotic concentrations. We systematically profiled Nile Red staining in *E. coli* and *P. aeruginosa* treated for 4 hours with ampicillin, cefepime, cefotaxime, meropenem, gentamicin, or ciprofloxacin. In both strains, fluorescence intensity increased within the MIC range defined by BMD, indicating that dye uptake serves as a reliable phenotypic indicator of susceptibility (Fig. 3A). While the overall staining trend was consistent between *E. coli* and *P. aeruginosa*, we observed that *P. aeruginosa* exhibited increased Nile Red-positive at slightly lower, sub-MIC antibiotic concentrations, particularly with β-lactams (Fig. S7A)

**Figure 3.**
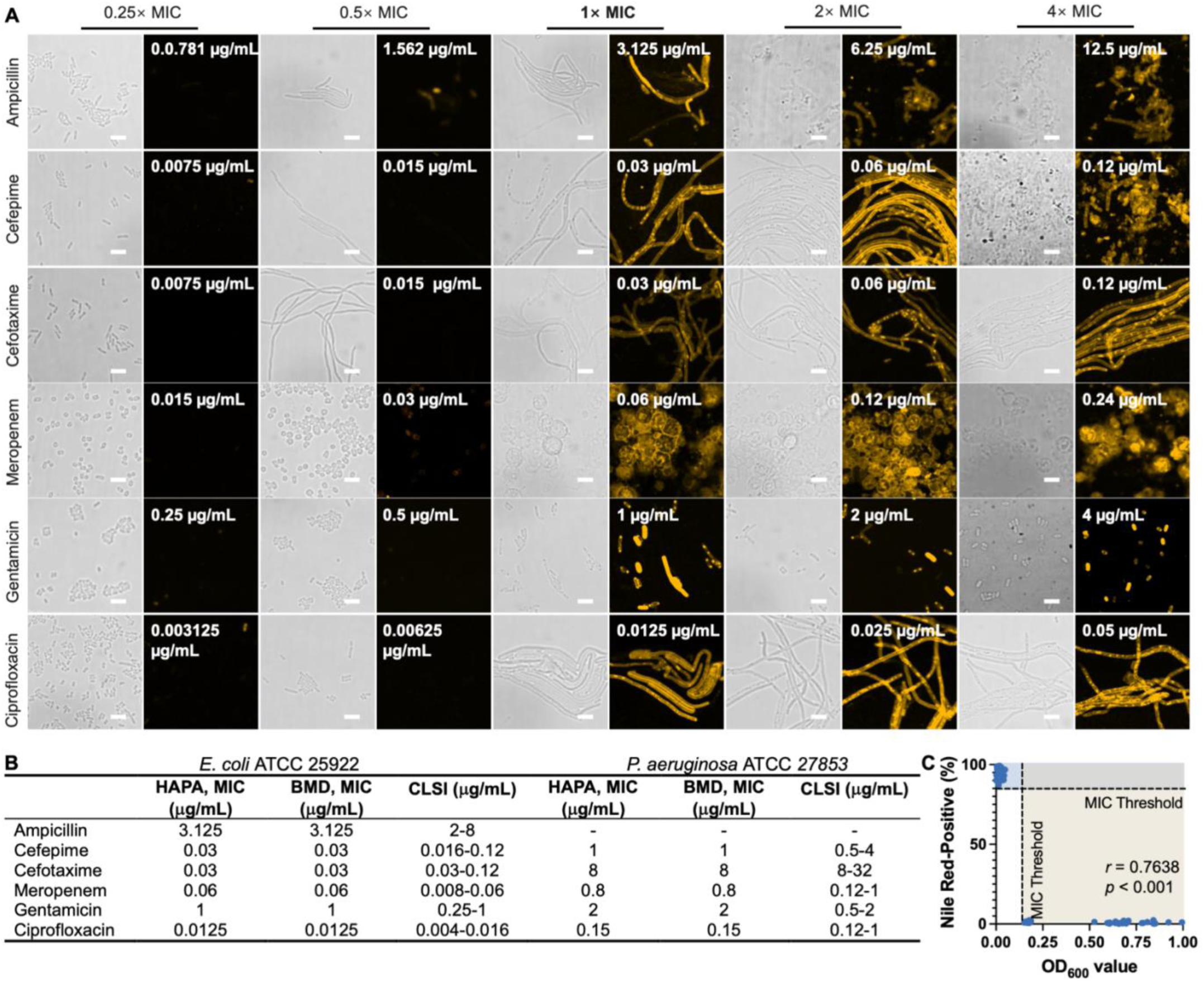
Phenotypic classification of antibiotic susceptibility based on fluorescence and morphology. **(A)** Representative images of *E. coli* treated with serial concentrations of ampicillin, cefepime, cefotaxime, meropenem, gentamicin, and ciprofloxacin for 4 hours. **(B)** Comparison of MIC values obtained by fluorescence-morphology phenotyping and BMD methods in *E. coli* ATCC 25922 and *P. aeruginosa* ATCC 27853, with CLSI-defined interpretive breakpoints shown for reference. **(C)** Correlation between OD_600_ and the proportion of Nile Red-positive cells across 60 antibiotic-treated conditions in *E. coli* ATCC 25922. Each point represents an individual measurement. The yellow-shaded region indicates the MIC threshold range defined by OD_600_ values from the BMD assay, while the blue-shaded region denotes the corresponding MIC threshold as determined by combined fluorescence and morphology-based phenotyping. Scale bars = 5 µm.

We reasoned that this early fluorescence response in *P. aeruginosa* at sub-MIC concentrations likely reflects underlying population heterogeneity.^25^ While a small subpopulation of sensitive cells is killed and becomes Nile Red-positive, the majority of cells remained filamentous and unstained (Fig. S7A). These unstained filamentous cells contributed substantially to OD_600_ and represented the surviving population, indicating that the antibiotic concentration was below the inhibitory threshold. To improve classification accuracy under these conditions, we incorporated antibiotic-induced morphological features as an additional phenotypic readout. For non-meropenem β-lactams, MIC-level responses were characterized by uniformly Nile Red-stained filamentous cells, whereas sub-MIC concentrations were associated with mixed populations, where only non-filamentous cells exhibited dye uptake and filamentous cells remained unstained. This composite phenotype allowed for more reliable discrimination of breakpoint responses.

To assess the accuracy of this combined phenotypic signature, we compared its susceptibility classifications against CLSI-defined breakpoints determined by BMD, using *E. coli* ATCC 25922 and *P. aeruginosa* ATCC 27853, the recommended quality control strains for AST standardization (Fig. 3B). A significant association between fluorescence response and bacterial growth inhibition was observed by correlating OD_600_ values with Nile Red-positive cell proportions after 16 and 4 hours of antibiotic exposure in *E. coli* ATCC 25922 (n = 60; Fig. 3C). Pearson correlation analysis revealed a statistically significant positive association (*r* = 0.7638, 95% CI: 0.6326–0.8524, *P* < 0.001), suggesting that the fluorescence signal broadly reflects changes in cell density. Notably, due to the binary-like transition in dye uptake once antibiotic concentrations approach the MIC, the actual association between bacterial viability and fluorescence intensity is likely stronger than indicated by the linear correlation coefficient.

These results prompted the development of the Hydrophobic Agent Permeability Assay (HAPA), a structured phenotypic AST method that integrates fluorescence and morphological responses at the single-cell level. HAPA enables interpretable, breakpoint-level susceptibility determination within 4 hours of antibiotic exposure, representing a 6-fold acceleration compared to the 24 hours required by conventional BMD methods (Fig. 4A).

**Figure 4.**
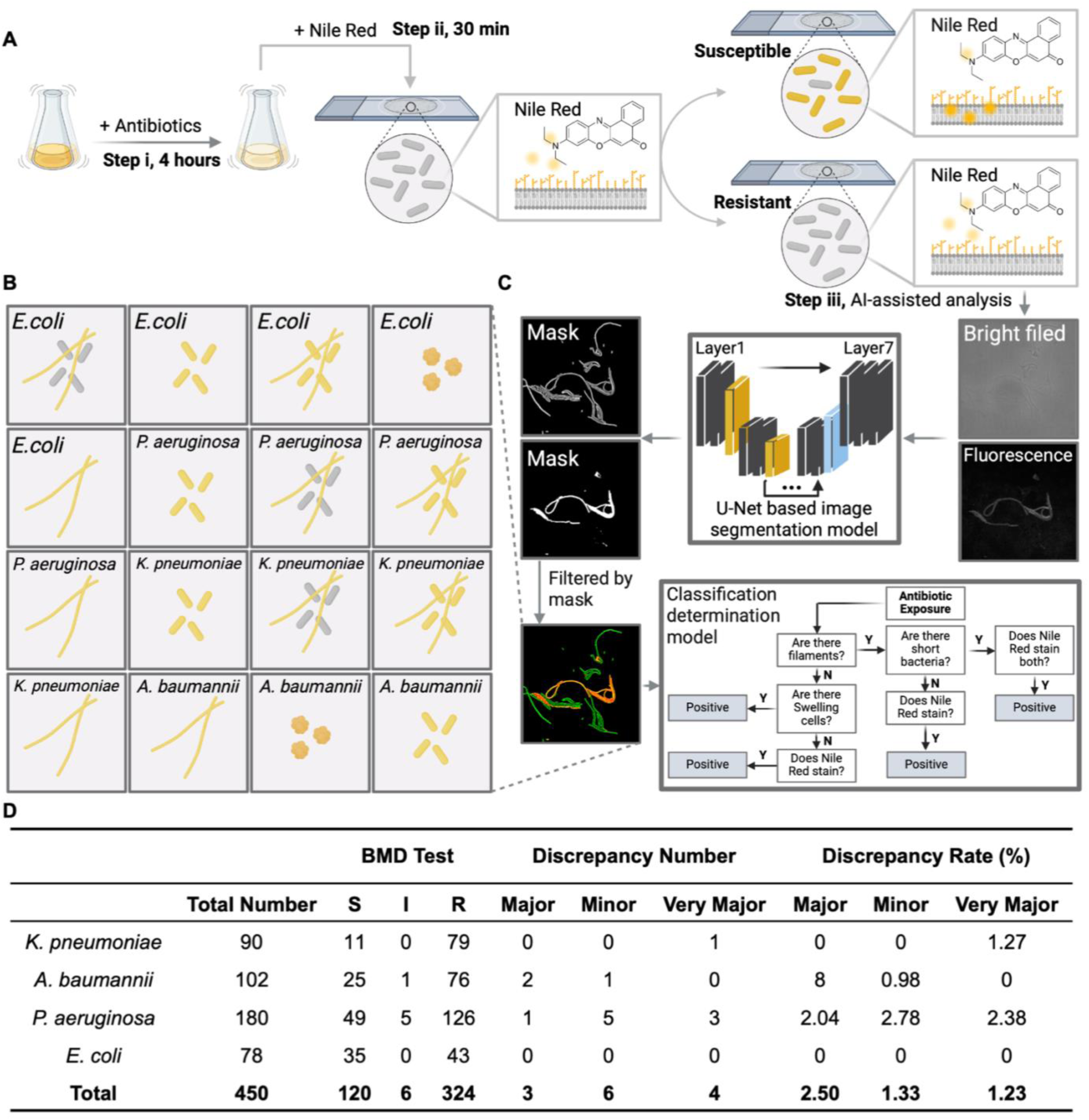
HAPA workflow, phenotypic templates, AI-assisted classification, and clinical performance. **(A)** Schematic overview of the HAPA. **(B)** MIC breakpoint-level phenotypic templates for four Gram-negative bacteria, based on combinations of fluorescence intensity and morphology. **(C)** Overview of the AI-assisted classification pipeline. **(D)** Summary of HAPA clinical performance across 456 antibiotic–strain tests using 81 Gram-negative clinical isolates.

To expand the applicability of HAPA, we extended the analysis to include two additional clinically relevant Gram-negative bacteria, *K. pneumoniae* 52145 and *A. baumannii* ATCC 19606. For each strain, we profiled Nile Red staining and morphological changes across a panel of antibiotic concentrations in susceptible reference strains, identifying distinct breakpoint-associated phenotypes (Fig. S8A; Fig. S9A).

Based on the identified breakpoint-level phenotypes across all four Gram-negative species, we compiled a curated reference image set to support the following machine learning-based MIC classification. For each antibiotic-strain pair, representative images at MIC concentrations (defined by BMD) were selected and annotated based on consistent morpho-fluorescent patterns. These images formed a ground truth template library used for supervised model training (Fig. 4B).

We further developed a machine learning framework designed to recognise integrated phenotypic patterns from image data (Fig. 4C). In the first stage, each input consisted of a co-registered pair of images: a phase-contrast image used to extract morphological features, and a corresponding fluorescence image used to quantify membrane permeability. These dual-channel images were processed by a U-Net-based segmentation model, which generated a mask delineating individual bacterial cells based on phase contrast and overlaid fluorescence signals. The segmentation accuracy assessment is shown in Fig. S10. Segmented and fused images were passed to a classification model trained to determine whether the input matched MIC breakpoint-level phenotypes. The determination of MIC in the HAPA method refers to the lowest antibiotic concentration at which a “positive” result is observed (Fig. 4C). This model referenced a curated phenotype template database and, upon identifying a breakpoint-level match, assigned a susceptibility category, susceptible, intermediate, or resistant, according to CLSI-defined MIC thresholds for the corresponding antibiotic concentration.

Finally, HAPA was applied to a panel of 75 clinical Gram-negative bacterial isolates, comprising *E. coli* (n = 13), *K. pneumoniae* (n = 15), *A. baumannii* (n = 17), and *P. aeruginosa* (n = 30). Each isolate was independently tested using both the HAPA assay and the standard BMD method under matched antibiotic conditions, including ampicillin, cefepime, cefotaxime, meropenem, gentamicin, and ciprofloxacin. For HAPA, susceptibility was determined based on fluorescence and morphological phenotyping at 4 hours post-treatment. Susceptibility classifications for both methods were assigned using CLSI-defined breakpoints, and diagnostic performance was assessed by comparing the categorical outcomes between HAPA and BMD according to FDA criteria for AST systems. As summarised in Figure 4D, the overall categorical agreement between HAPA and BMD was 97.11%, exceeding FDA-recommended thresholds (>90%). The observed minor error (mE), major error (ME), and very major error (VME) rates were 1.33%, 2.50%, and 1.23%, respectively, indicating strong concordance and clinical applicability of HAPA across diverse Gram-negative species. Moreover, the receiver operating characteristic (ROC) curve for all tested bacterial species was generated, using BMD-determined breakpoints as the ground truth. The area under the curve (AUC) values exceeded 0.956 across all species (*P* < 0.001) (Fig. S11A), confirming the high sensitivity and specificity of the HAPA classification algorithm. These findings underscore the translational potential of HAPA for rapid, high-throughput AST in routine microbiology laboratories.

## Discussion

The increasing global burden of antimicrobial resistance has created an urgent demand for rapid, reliable, and clinically deployable AST strategies. In this study, we developed and validated a novel phenotypic platform, HAPA, which enables MIC estimation within 4 hours by detecting antibiotic-induced changes in membrane permeability and morphology using Nile Red staining and automated image analysis. This method represents a significant advancement toward rapid, microscopy-based AST capable of single-cell resolution and interpretability.

In this study, our findings highlight a previously underappreciated mechanism by which bactericidal antibiotics modulate bacterial envelope properties. While the outer membrane of Gram-negative bacteria is traditionally viewed as a static physical barrier, our data suggest that oxidative stress triggered by antibiotics with diverse intracellular targets can remodel outer membrane permeability in a ROS-dependent manner. Notably, this process enhances access of hydrophobic agents such as Nile Red without compromising membrane integrity, distinguishing it from classic lytic mechanisms.^23^ These results extend the role of ROS beyond canonical intracellular damage, such as DNA damage and protein oxidation, to include outer membrane lipid alterations, suggesting a new layer of antibiotic-induced stress response. This mechanistic insight provides a foundation for leveraging fluorescent agents’ permeability as an early and functional biomarker of antibiotic activity.

Building on this mechanistic understanding, we demonstrate that ROS-associated fluorescence responses can serve as quantitative phenotypic indicators of susceptibility. In both reference and clinical strains, susceptible bacteria exhibited antibiotic concentration-dependent increases in hydrophobic agents’ uptake and morphological changes, which consistently aligned with MIC thresholds defined by BMD. Unlike conventional growth-based methods that capture endpoint viability, our approach offers a more immediate snapshot of bacterial stress response, enabling rapid phenotypic stratification. The ability to detect susceptibility-related phenotypes within hours is particularly valuable in the context of mixed or partially resistant populations, where bulk OD_600_-based methods may obscure early subpopulation responses. These features position HAPA as a scalable tool for accelerated AST with improved temporal resolution.

A key advantage of HAPA lies in its ability to resolve phenotypic heterogeneity within bacterial populations, which is an often-overlooked challenge in phenotypic AST. Previous methods such as single cell morphology assay (SCMA),^26^ which rely solely on morphological changes, may misclassify susceptibility in strains exhibiting mixed populations. For instance, in *P. aeruginosa*, sub-MIC β-lactam exposure frequently produces coexisting short and filamentous cells. In SCMA, such morphological heterogeneity is often interpreted as indicative of resistance.^26^ However, our data reveal that fluorescence staining provides a critical discriminator: if filamentous cells remain unstained while only short cells uptake Nile Red, the population remains below the MIC; conversely, uniformly stained filaments mark the true inhibitory threshold. This refined interpretation prevents misclassification and enables more precise MIC estimation.

Although the current implementation of HAPA employed a spinning disk confocal system, the imaging requirements are minimal. Since HAPA relies primarily on bacterial morphology and fluorescence intensity, rather than subcellular structures or high-resolution localisation, it is fully compatible with lower-cost, widefield or LED-based fluorescence imaging systems. This imaging flexibility enhances the feasibility of HAPA for use in decentralised laboratories or low-resource settings.

Moreover, the use of hydrophobic agents in AST has traditionally relied on chemical fixation,^27^ which limits temporal resolution and distorts native membrane physiology. In contrast, HAPA captures dynamic, treatment-induced permeability shifts, allowing functional resolution of MIC breakpoints within hours. This real-time capability represents a significant methodological advance, especially for applications requiring rapid susceptibility readouts.

Finally, to facilitate high-throughput implementation and standardised interpretation, the HAPA assay was integrated with a machine learning framework capable of automated breakpoint classification, further supporting its potential for deployment in clinical or point-of-care settings.

Despite these promising results, several limitations and future development directions should be acknowledged. First, although our findings support a model in which ROS-mediated lipid oxidation enhances membrane permeability, direct biochemical evidence, such as structural characterisation of oxidised LPS species, needs to be convinced. Second, integration of HAPA into fully automated platforms remains a critical step toward the real-world application. Future efforts may focus on microfluidic system design to minimise sample handling, such as eliminating centrifugation and manual preparation steps prior to imaging, thereby further reducing assay time. Third, while this study evaluated six representative antibiotics across four Gram-negative species, broader validation across a wider antibiotic spectrum will be essential to establish HAPA as a comprehensive AST platform. Lastly, future work will also aim to validate the HAPA platform directly on clinical specimens such as blood or urine, with multicentre studies currently underway to assess its diagnostic performance in real-world infection settings.

In conclusion, this study reveals a ROS-dependent mechanism by which antibiotic exposure alters outer membrane permeability in Gram-negative bacteria and leverages this insight to establish a rapid, interpretable phenotypic AST platform. HAPA has the potential to reduce AST turnaround time, support antimicrobial stewardship, and serve as a valuable complement to culture-based and molecular diagnostics in both centralised laboratories and point-of-care settings.

## Data availability statement

All data supporting the findings of this study are available within the paper and its Supplementary Information.

## Contributors

L.Z. contributed the initial experimental idea. L.Z., J.Z., L.J., and D.J. designed the experiments. J.L. and A.C provided microbiological oversight of clinical isolated samples. G.P., L.Z., and D.L. conducted experiments and analysed data. L.Z., and D.J. wrote the manuscript with input from all co-authors.

## Data Availability

All data produced in the present study are available upon reasonable request to the authors

## Acknowledgements

This work was supported by the Australian Research Council (ARC) Laureate Fellowship (FL210100180), ARC Future Fellowships (FT220100018 and FT220100152) and ARC Centre of Excellence in Quantum Biotechnology (CE230100021). J.L. is an Australian NHMRC Investigator Fellow (APP2025937). We thank Dr Ji Lu from the Australian Centre for Water and Environmental Biotechnology (ACWEB), University of Queensland, Australia, for generously providing clinically isolated *K. pneumoniae* strains. We also acknowledge Prof. Jon Iredell and Dr. Carola Venturini from the Westmead Institute for Medical Research, Westmead Hospital, Australia, Ms. Heidi Yu and Mr. Ke (Kenny) Chen from the Department of Microbiology, Biomedicine Discovery Institute, Monash University, Australia, and Dr Liping Li from the School of Natural Sciences, Macquarie University, Australia, for their support in providing the clinical isolates.

---

**Figure S1.**
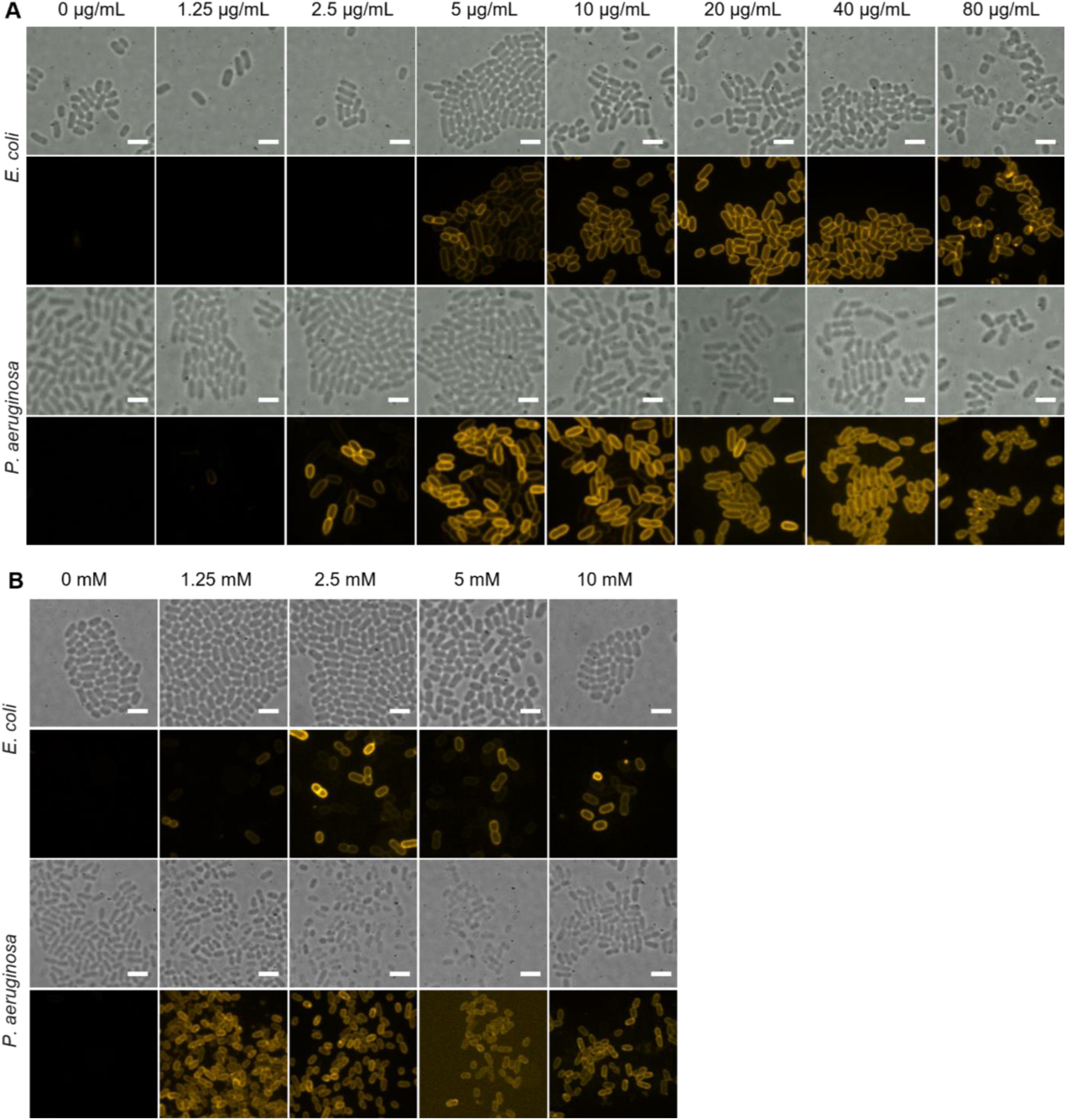
Dose-dependent enhancement of Nile Red uptake by PMB and EDTA in Gram-negative bacteria. **(A)** Representative bright field and fluorescence images of *E. coli* and *P. aeruginosa* treated with increasing concentrations of PMB for 5 minutes, followed by staining with Nile Red. **(B)** Corresponding images of *E. coli* and *P. aeruginosa* treated with increasing concentrations of EDTA for 0.5 hours, followed by staining with Nile Red. Scale bars = 2 µm.

**Figure S2.**
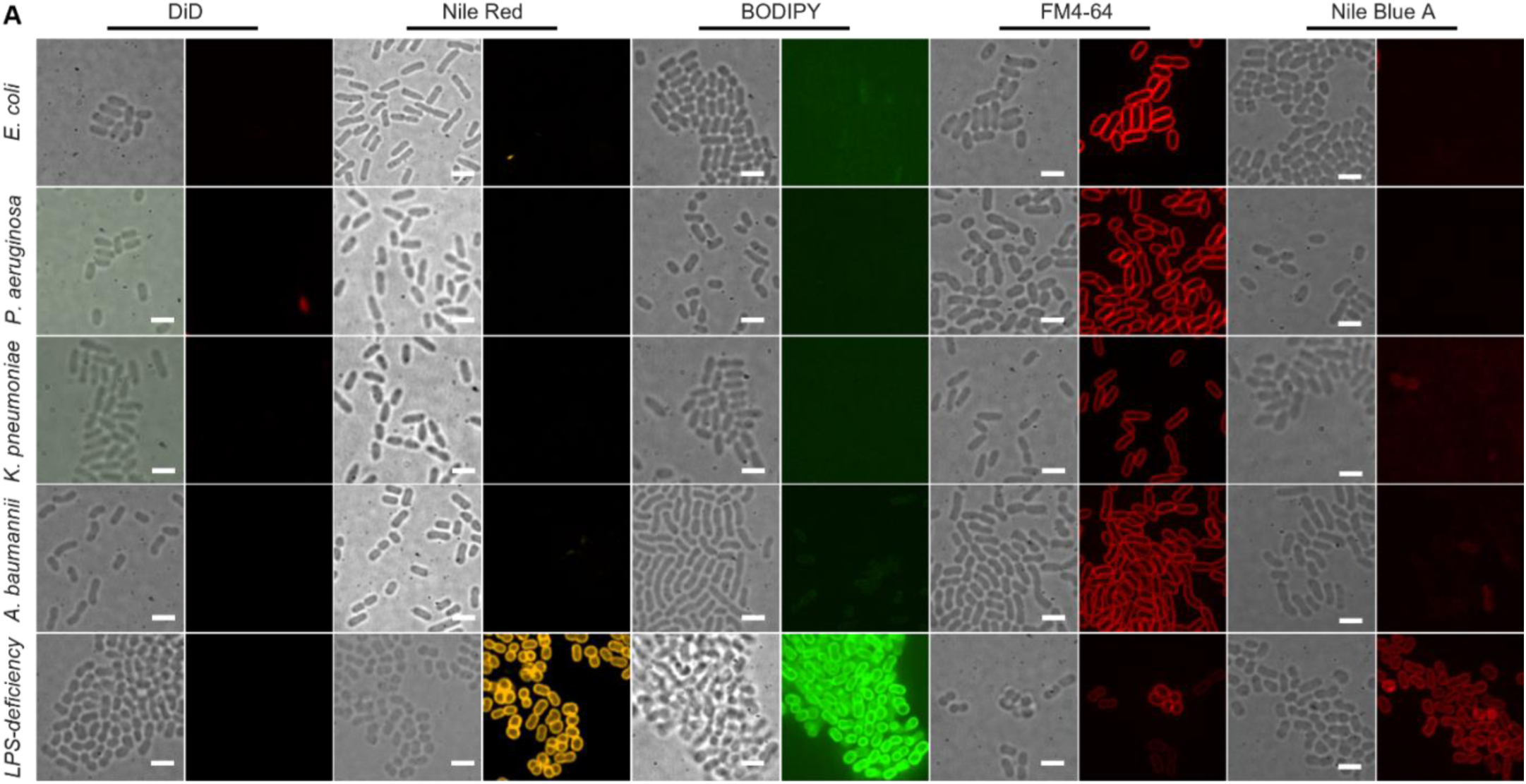
Selective exclusion of hydrophobic dyes by the outer membrane of Gram-negative bacteria. **(A)** Representative bright-field and fluorescence images of *E. coli*, *P. aeruginosa*, *K. pneumoniae*, and *A. baumannii* wild-type strains, alongside an LPS-deficient mutant of *A. baumannii*, stained with five membrane dyes: DiD, Nile Red, BODIPY, FM 4-64, and Nile Blue A. Scale bars = 2 µm.

**Figure S3.**
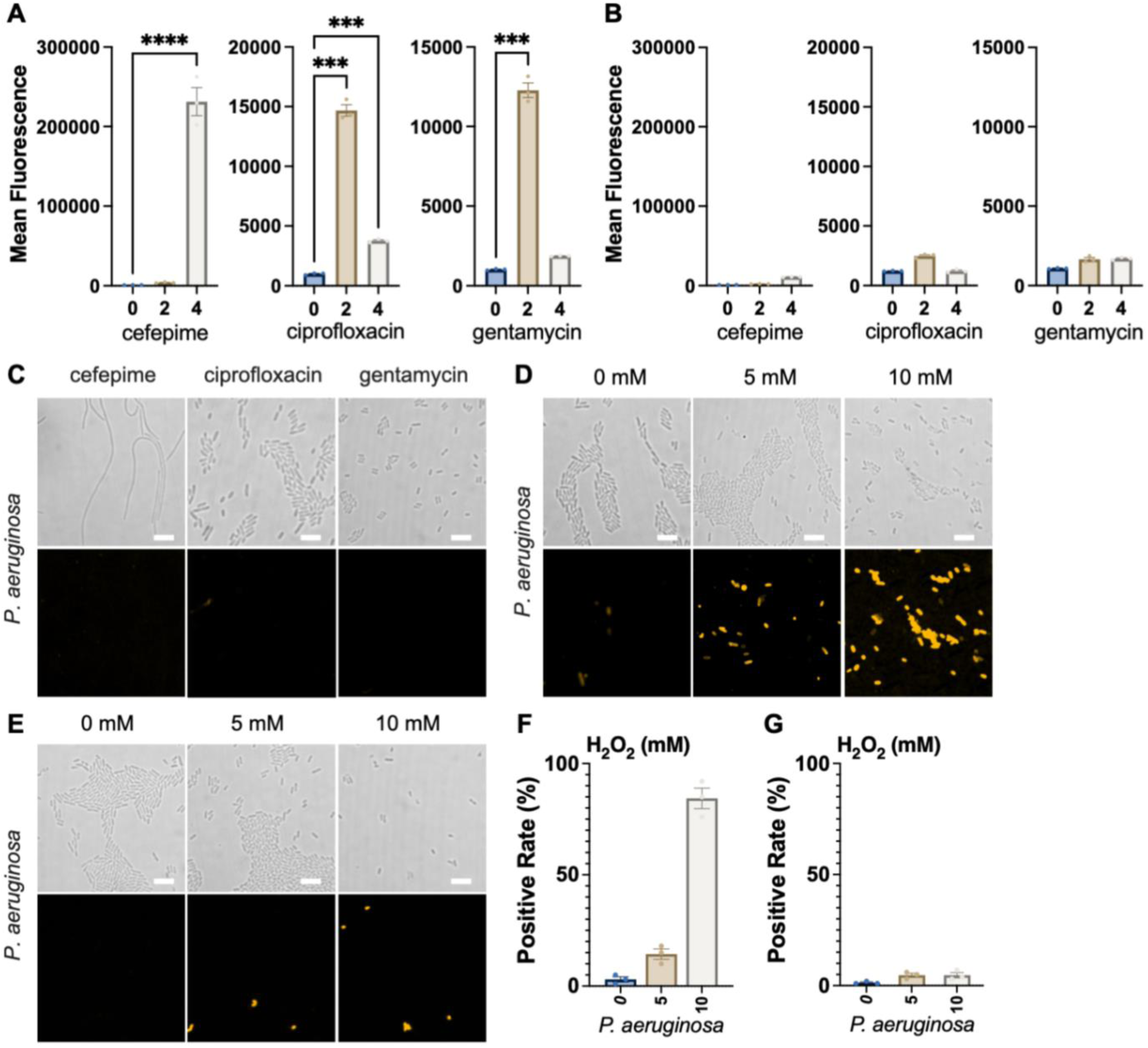
Antibiotic-induced oxidative stress enhances membrane permeability in *P. aeruginosa*. **(A)** Intracellular ROS levels measured after antibiotic treatment in *P. aeruginosa* ATCC 47085 treated with antibiotics over 0, 2, and 4 hours. **(B)** Co-treatment with GSH (50 mM) abolished antibiotic-induced ROS production in *P. aeruginosa* ATCC 47085. **(C)** Representative fluorescence and bright-field images of *P. aeruginosa* ATCC 47085 co-treated with antibiotic (cefepime at 0.03 µg/mL, ciprofloxacin at 0.0125 µg/mL, or gentamycin at 1 µg/mL) and GSH (50 mM) for 4 hours. **(D)** Exogenous ROS application via H₂O₂ treatment (0, 5, 10 mM, 1 hour) induced dose-dependent increases in Nile Red fluorescence in *P. aeruginosa* ATCC 47085. **(E)** PI staining following H₂O₂ exposure (0, 5, 10 mM, 1 hour) did not show increased signal in *P. aeruginosa* ATCC 47085 **(F and G)** Percentage of Nile Red-positive and PI-positive cells in *P. aeruginosa* ATCC 47085 after H₂O₂ treatment. The positive rate was defined as the proportion of Nile Red- or PI-stained cells relative to the total number of cells within a single image frame. Quantification of fluorescence intensity is shown as mean ± SEM from three independent experiments. Statistical significance determined by one-way ANOVA; *, *P* < 0.05; **, *P* < 0.01; ***, *P* < 0.001. Scale bars, 5 µm.

**Figure S4.**
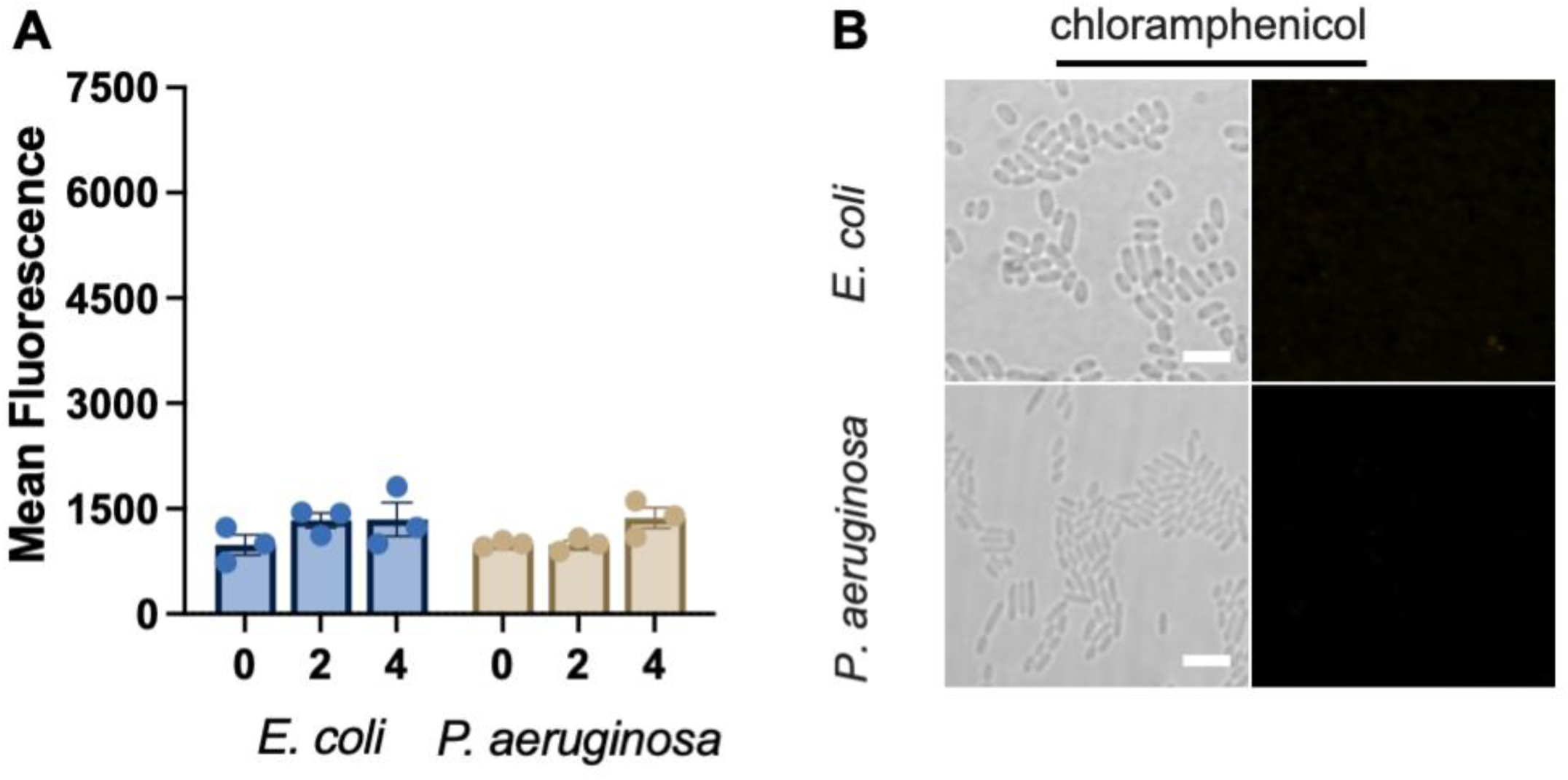
Chloramphenicol fails to enhance the ROS generation and Nile Red uptake. **(A)** Intracellular ROS levels measured after chloramphenicol treatment in *E. coli* at 12.5 μg/mL or *P. aeruginosa* at 50 μg/mL for 0, 2, and 4 hours. **(B)** Representative bright-field and fluorescence images of the *E. coli* or *P. aeruginosa* at 12.5 or 50 μg/mL for 4 hours, followed by staining with Nile Red. Quantification of fluorescence intensity is shown as mean ± SEM from three independent experiments. Scale bars = 2 µm.

**Figure S5.**
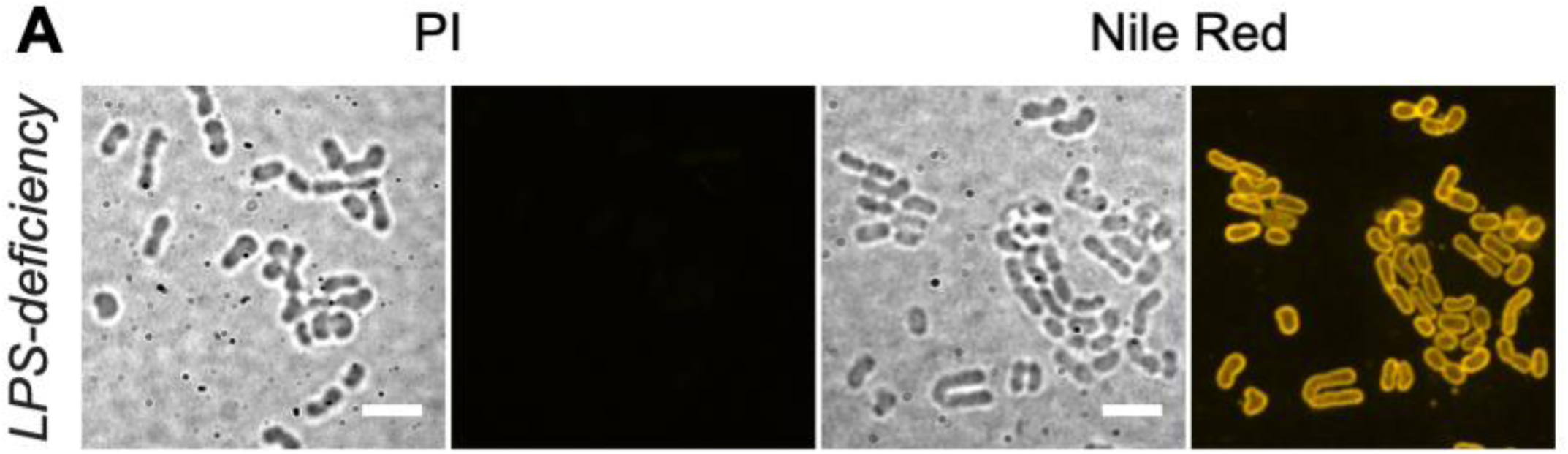
PI fails to stain LPS-deficient *A. baumannii*, despite enhanced Nile Red uptake. **(A)** Representative bright-field and fluorescence images of LPS-deficient *A. baumannii* stained with PI at 10 μg/mL, or Nile Red at 2.5 μg/mL for 30 minutes. Scale bars = 2 µm.

**Figure S6.**
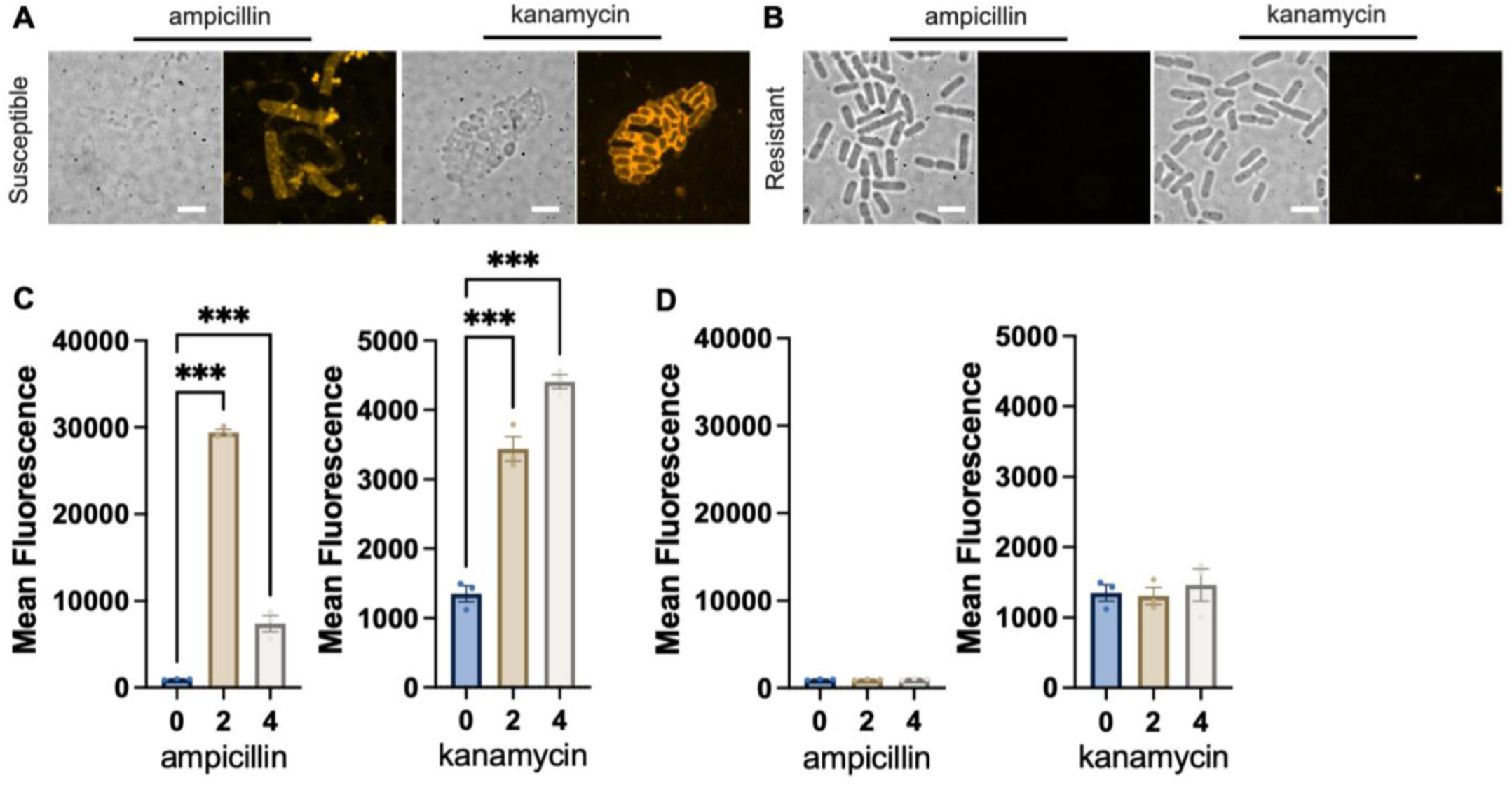
Antibiotic-induced ROS generation and Nile Red uptake are absent in resistant *E. coli* strains. **(A)** Representative bright-field and fluorescence images of drug-susceptible *E. coli* after 4-hour exposure to ampicillin at 6.25 µg/mL or kanamycin at 12.5 µg/mL. **(B)** Representative bright-field and fluorescence images of *E. coli* carrying plasmids encoding resistance to ampicillin or kanamycin after 4-hour exposure to ampicillin at 6.25 µg/mL or kanamycin at 12.5 µg/mL. **(C and D)** Intracellular ROS levels measured in susceptible (C) and resistant (D) *E. coli* strains after 0, 2, or 4 hours of ampicillin (6.25 µg/mL) and kanamycin (12.5 µg/mL) treatment. Quantification of fluorescence intensity is shown as mean ± SEM from three independent experiments. Statistical significance determined by one-way ANOVA; ***, *P* < 0.001. Scale bars = 2 µm.

**Figure S7.**
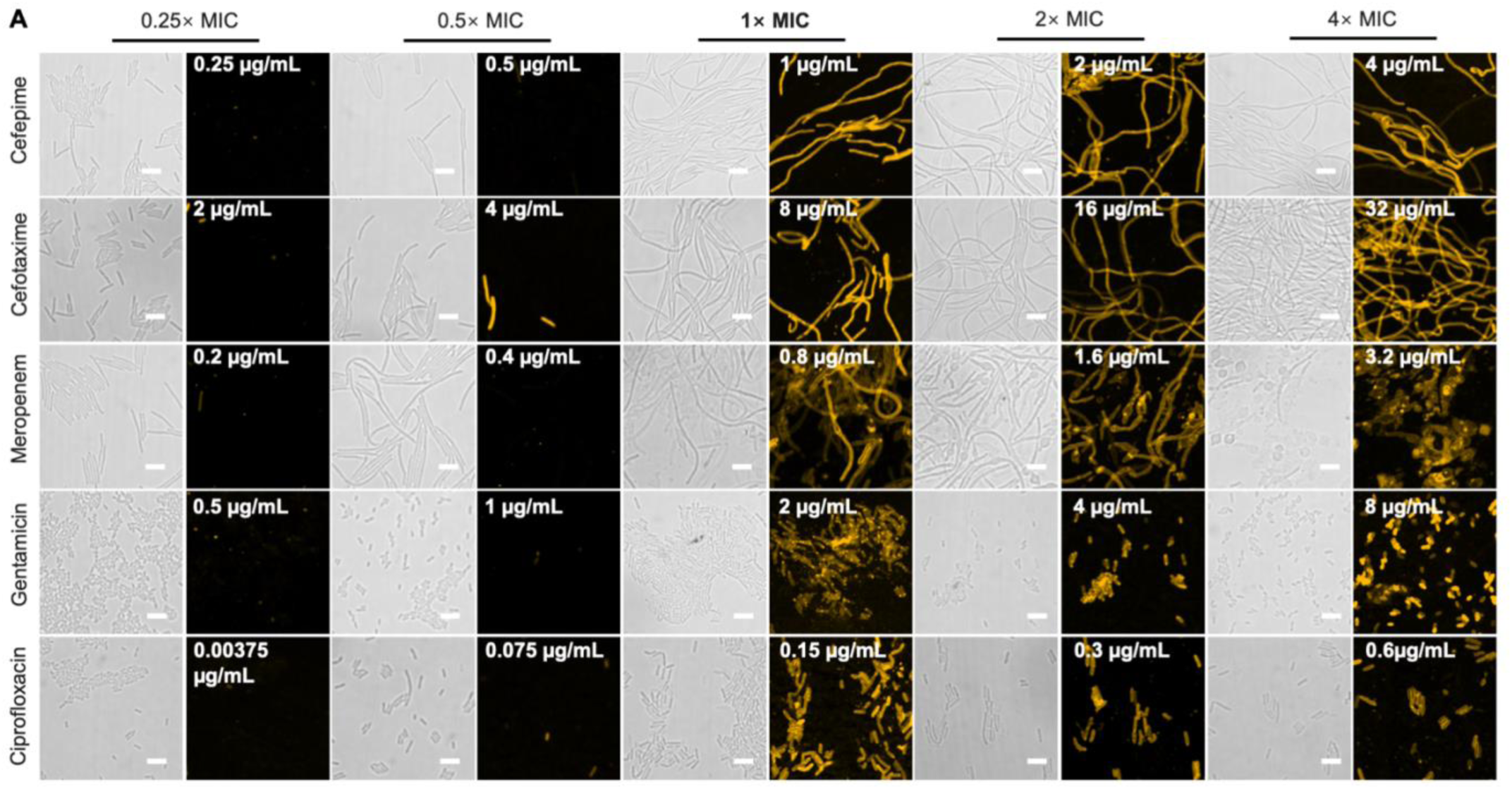
Phenotypic classification of antibiotic susceptibility based on fluorescence and morphology in *P. aeruginosa*. **(A)** Representative images of *P. aeruginosa* ATCC 27853 treated with serial concentrations of cefepime, cefotaxime, meropenem, gentamicin, and ciprofloxacin for 4 hours. Scale bars = 5 µm.

**Figure S8.**
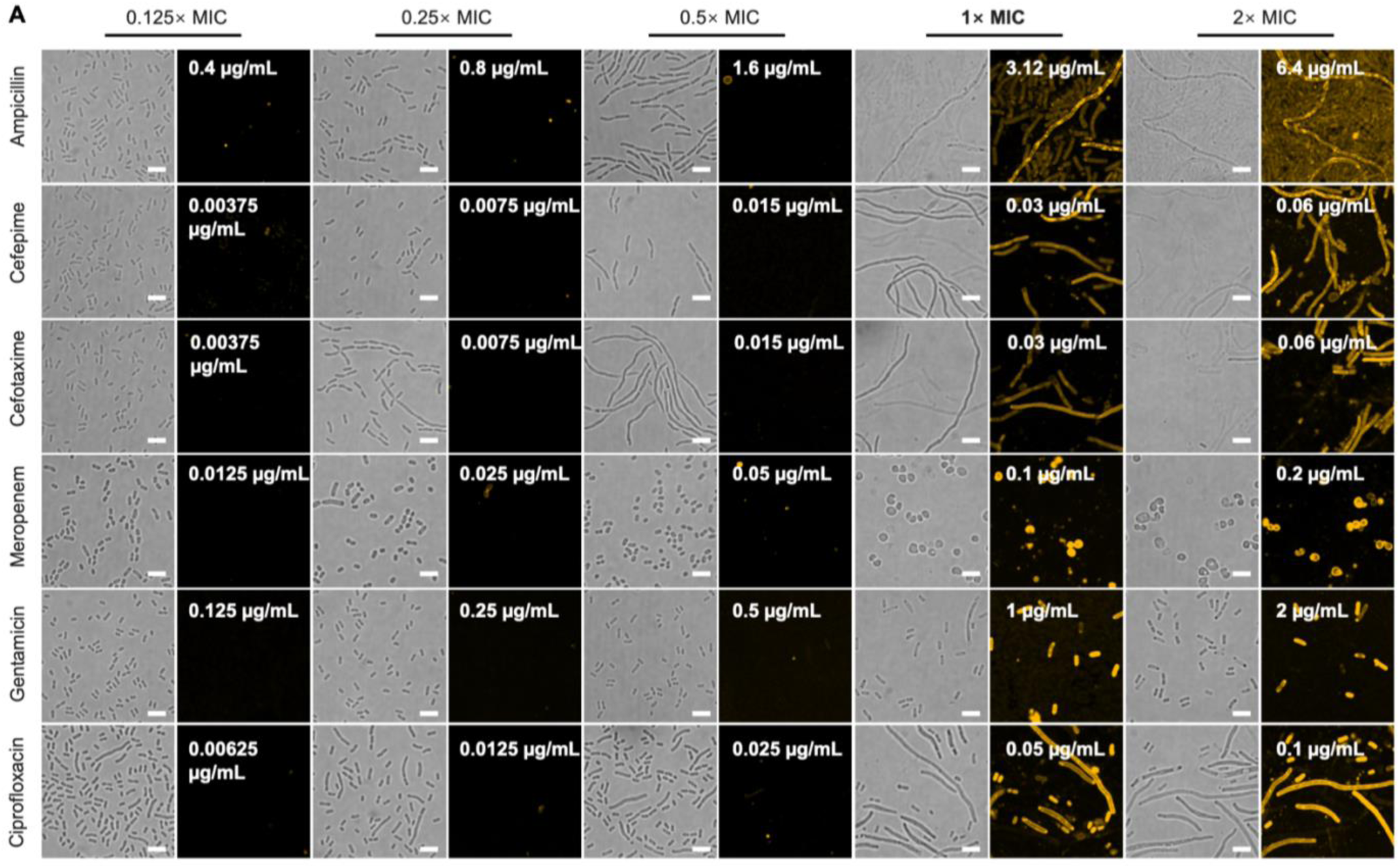
Phenotypic classification of antibiotic susceptibility based on fluorescence and morphology in *K. pneumoniae*. **(A)** Representative images of *K. pneumoniae* ATCC 52145 treated with serial concentrations of ampicillin, cefepime, cefotaxime, meropenem, gentamicin, and ciprofloxacin for 4 hours. Scale bars = 5 µm.

**Figure S9.**
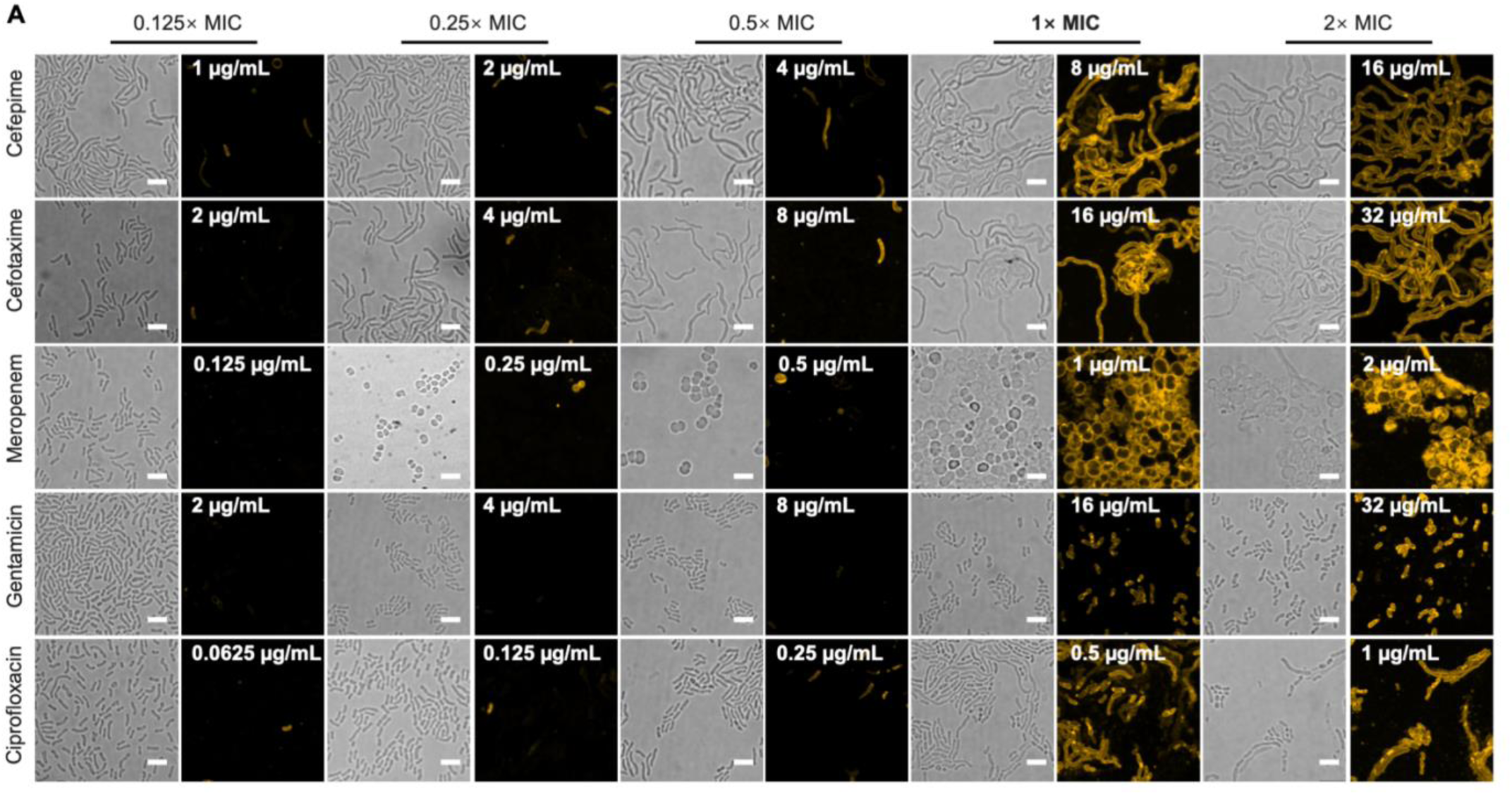
Phenotypic classification of antibiotic susceptibility based on fluorescence and morphology in *A. baumannii*. **(A)** Representative images of *A. baumannii* 19606 treated with serial concentrations of cefepime, cefotaxime, meropenem, gentamicin, and ciprofloxacin for 4 hours. Scale bars = 5 µm.

**Figure S10.**
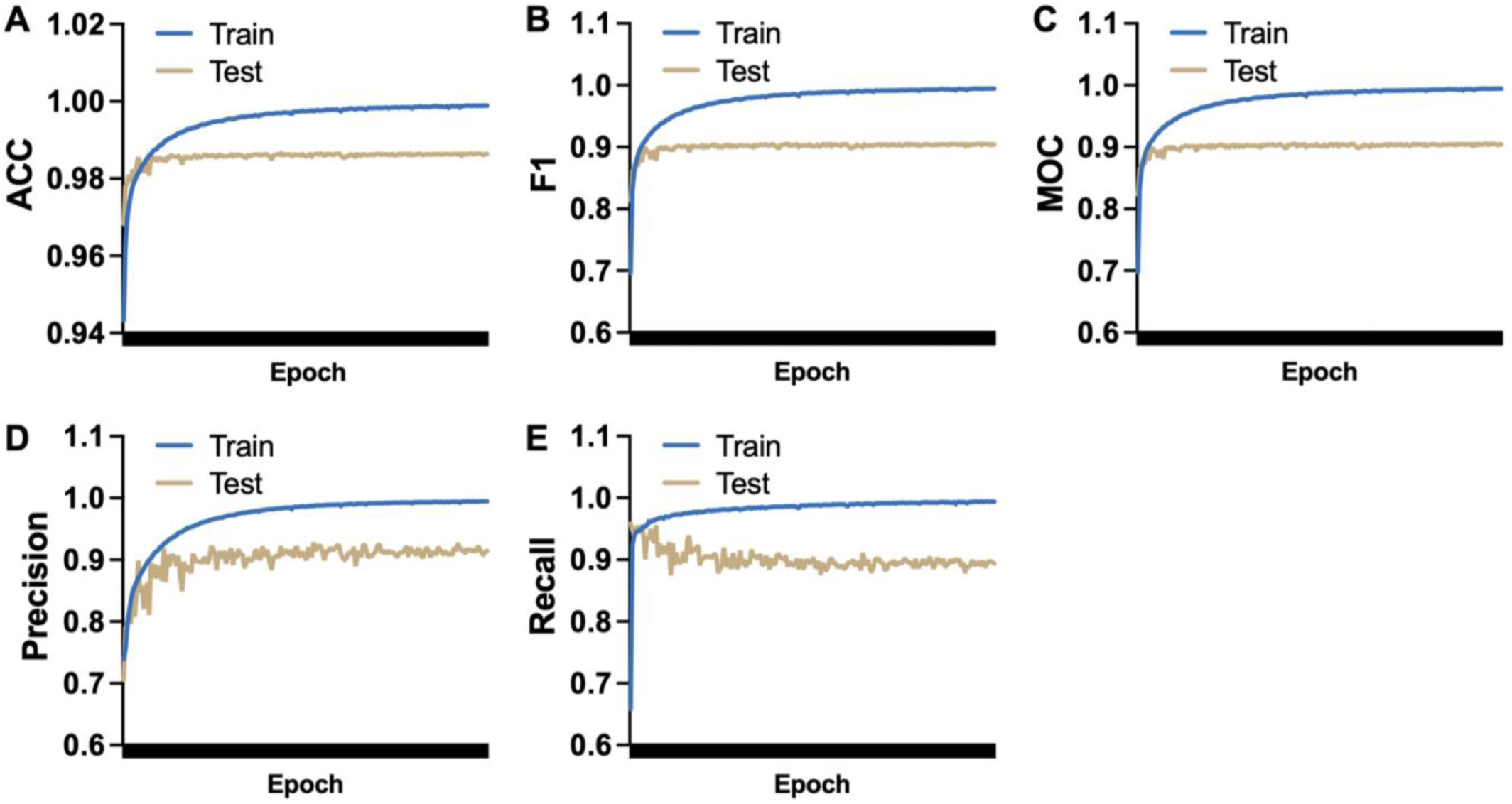
Quantitative evaluation of U-Net segmentation performance across training epochs. Segmentation performance was assessed by five metrics: **(A)** Accuracy (ACC). **(B)** F1 score. **(C)** Mean overlap coefficient (MOC). **(D)** Precision. **(E)** Recall. Curves represent metric values across training (blue) and testing (gold) datasets over 200 epochs. All metrics on the test set exceeded 90%. The gap between training and testing curves reflects expected generalization limits without overfitting.

**Figure S11.**
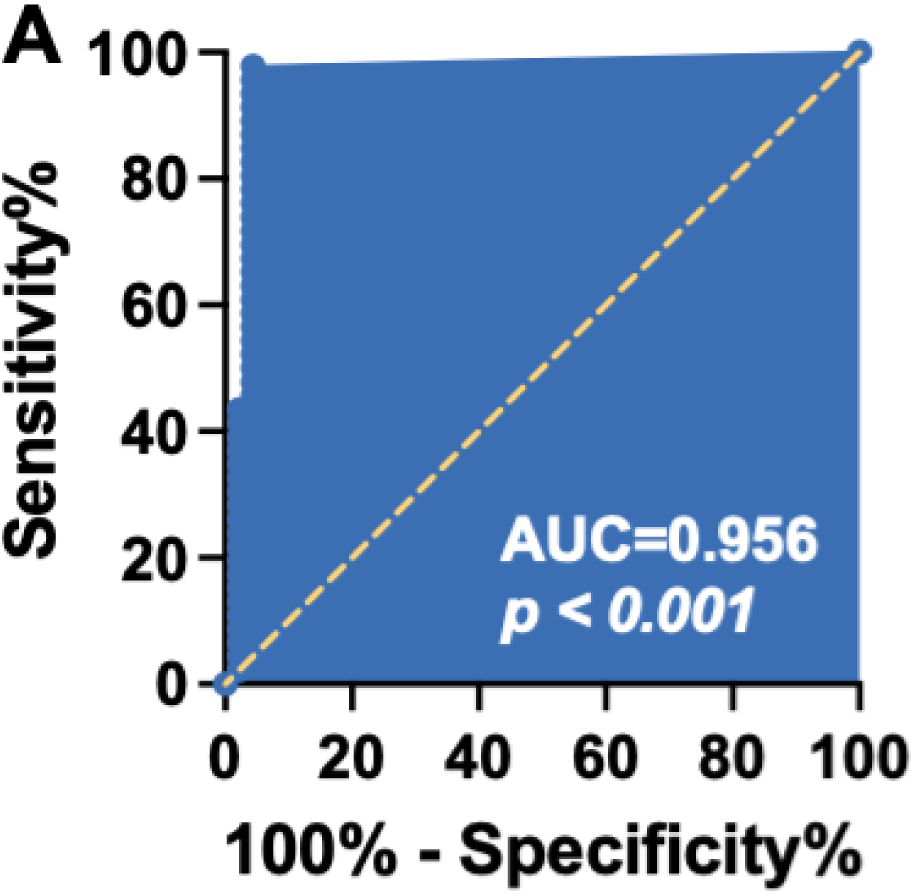
ROC curve evaluating the performance of the HAPA in classifying antibiotic susceptibility. **(A)** ROC curve of the HAPA method compared to the CLSI reference standard for binary classification (susceptible vs resistant).

